# Axonal Injury, Sleep Disturbances, and Memory following Traumatic Brain Injury

**DOI:** 10.1101/2024.03.05.24303449

**Authors:** Emma M. Tinney, Goretti España-Irla, Aaron E.L. Warren, Lauren N. Whitehurst, Alexandra M Stillman, Charles H. Hillman, Timothy P. Morris, Alzheimer’s Disease Neuroimaging Initiative

**Author notes:** Data used in preparation of this article were obtained from the Alzheimer’s Disease Neuroimaging Initiative (ADNI) database (adni.loni.usc.edu). As such, the investigators within the ADNI contributed to the design and implementation of ADNI and/or provided data but did not participate in analysis or writing of this report. A complete listing of ADNI investigators can be found at: http://adni.loni.usc.edu/wp-content/uploads/how_to_apply/ADNI_Acknowledgement_List.pdf. Corresponding author: Timothy P. Morris, PhD. Center for Cognitive and Brain Health, Northeastern University, 805 Columbus Ave, Boston, USA, 02115.

## Abstract

**Objectives:** Traumatic brain injury (TBI) is associated with sleep deficits, but it is not clear why some report sleep disturbances and others do not. The objective of this study was to assess the associations between axonal injury, sleep, and memory in chronic and acute TBI.

**Methods:** Data were acquired from two independent datasets which included 156 older adult veterans (69.8 years) from the Alzheimer’s Disease Neuroimaging Initiative (ADNI) with prior moderate-severe TBIs and 90 (69.2 years) without a TBI and 374 participants (39.6 years) from Transforming Research and Clinical Knowledge in TBI (TRACK-TBI) with a recent mild TBI (mTBI) and 87 controls (39.6 years), all who completed an MRI, memory assessment, and sleep questionnaire.

**Results:** Older adults with a prior TBI had a significant association between axial diffusivity in the left anterior internal capsule (ALIC) and sleep disturbances [95% CI(5.0e+07, 1.7e+08), p ≤ .01]. This association was significantly different [95% CI(6.8e+07,2.2e+08), p=.01] from controls. ALIC predicted changes in memory over one-year in TBI [95% CI(−1.8e+08,-2.7e+07), p=.03]. We externally validated those findings in TRACK-TBI where ALIC axial diffusivity within two-weeks after injury was significantly associated with higher sleep disturbances in the TBI group at two-weeks [95% CI(−7.2e-06, −1.9e-04), p=0.04], six-months [95% CI(−4.2e-06,-1.3e-04), p≤ .01] and 12-months post-injury [95% CI(−5.2e-06, −1.2e-04), p=0.03]. These associations not seen in controls.

**Interpretations:** Axonal injury to the ALIC is robustly associated with sleep disturbances in multiple TBI populations. Early assessment of ALIC damage following mTBI could identify those at risk for persistent sleep disturbances following injury.

## Introduction

Traumatic brain injury (TBI) has the highest incidence of all common neurological disorders^1^ and patients report a variety of behavioral symptoms that can last for years post injury. However, it is not clear why some individuals report sleep disturbances and others do not. Around half of all adults with TBI report sleep disturbances including insomnia, hypersomnia, and poor sleep quality^2^, with sleep disorders being three times more common in TBI patients than in the general population^3^.

Importantly, chronic sleep disorders are known to have negative effects on memory function across the lifespan ^4^. Studies also show that a history of TBI can be associated with increased risk of cognitive decline ^6^ and pathological aging ^7^. In non-TBI populations, sleep disturbances have been previously associated with the integrity of white matter in specific tracts in the brain. For example, decreased white matter integrity of association fibers such as the superior longitudinal fasciculi, commissarial fibers in the corpus callosum, and projection fibers of the internal capsule and corona radiata have been reported in non-injured adults with poor sleep quality ^8^ and insomnia ^9^. A hallmark of TBI is diffuse axonal injury (DAI) caused by shear and tensile forces placed upon the brain during the initial impact, where almost every tract studied shows axonal injury following acute mTBI^10^. Importantly, there is evidence that white matter lesions caused by disease or other injury types lead to sleep disturbances^11^. For example, demyelination in patients with multiple sclerosis often precipitates sleep deficits, with the precise location of the lesions determining the type of deficit ^11^. Nevertheless, the relationship between sleep and white matter is complex and recent work has proposed that the relationship between structural alterations and sleep is likely bi-directional ^12,13^.

Identifying mechanisms of sleep disturbances in TBI is important for several reasons. First, given poor sleep quality is linked with cognitive decline, identifying mechanisms of sleep disturbances after injury can improve clinically targeted interventions. Second, it can increase insights into why some individuals with TBI report sleep disturbances and others do not. Third, it can help refine prognostic models designed to predict behavioral outcomes in those living with the consequences of injury.

Here, we use diffusion-weighted imaging to test the associations between axonal injury to a-priori sleep-related white matter tracts and sleep disturbances in older adults who suffered a TBI decades prior and adults with an acute (< two weeks) mild TBI. We test these associations in two independent cohorts and compare them to non-injured adults to test if they are specific to TBI. We further test whether sleep-related axonal injury predicts memory changes over one-year in both cohorts.

## Methods

### Participants

#### Older adults with TBI history: ADNI/DoD

Data from the Brain Aging in Vietnam War Veterans (ADNI/DoD) dataset, part of the Alzheimer’s Disease Neuroimaging Initiative (ADNI) database were used. Full details of ADNI/DoD and participant inclusion criteria is found in supplementary materials 1. For the current study, 246 male veterans aged 60-85 with (N=156) and without a history (N = 90) of non-penetrating moderate-severe TBI, who had complete diffusion-weighted MRI scans and Pittsburgh sleep quality index (PSQI) scores were included. Session one consisted of a memory assessment (Ray Auditory Verbal Learning Test (RAVLT)), magnetic resonance imaging (MRI) scan, and self-report sleep assessment. All measurements were repeated one year later. Participant characteristics for ADNI/DoD are in Table 1.

**Table 1:** ADNIDOD Participant Characteristics.

|  | **TBI (mean ± SD)** | **No TBI (mean ± SD)** | **Difference** |
| --- | --- | --- | --- |
| Age | 69.85 ± 4.6 | 69.2 ± 4.7 | t=1.25 |
| Sex | All Male | All Male | X<sup>2</sup>=0 |
| Years of Education | 15.21 ± 2.4 | 15.49 ± 2.2 | t=-0.92 |
| American Indian (%) | 1% | 1% | X<sup>2</sup>=0 |
| Asian (%) | 0% | 2% | X<sup>2</sup>=1.02 |
| Black/African American (%) | 9% | 8% | X<sup>2</sup>=0 |
| White (%) | 93% | 81% | X<sup>2</sup>=0.96 |
| More than 1/Unknown (%) | 3% | 7% | X<sup>2</sup>=0.12 |
| Depression<sup>a</sup> (n) | 39 | 22 | X<sup>2</sup>=0.27 |
| PTSD Current<sup>b</sup> (n) | 61 | 41 | X<sup>2</sup><0.01 |
| PTSD Lifetime<sup>c</sup> (n) | 85 | 45 | X<sup>2</sup>=2.48 |
| MMSE<sup>d</sup> | 28.15 ± 1.8 | 28.3 ± 1.6 | t=-0.81 |
| Time Since Injury (years) | 25 ± 14.9 | NA | NA |
| Total number of TBIs | 1.6 ± 1.3 | NA | NA |
| **Pittsburg Sleep Quality Index** | 11.76 ± 2.1 | 11.63 ± 2.6 | t=0.81 |
| Subjective Sleep Quality | 1.22 ± 0.78 | 1.25 ± 0.72 | t=-0.26 |
| Sleep Latency | 1.22 ± 1.07 | 1.14 ± 0.95 | t=0.63 |
| Sleep Duration | 1.63 ± 1.11 | 1.72 ± 1.05 | t=-0.61 |
| Sleep Efficiency | 2.82 ± 0.65 | 2.91 ± 0.36 | t=-1.05 |
| Sleep Disturbance | 2.56 ± 0.49 | 2.42 ± 0.54 | t=2.02 |
| Use of Sleep medication | 2.24 ± 1.14 | 2.19 ± 1.22 | t=0.27 |
| Daytime Dysfunction | 1.32 ± 0.56 | 1.31 ± 0.69 | t=0.13 |
| APOE<sup>e</sup> (n) | 43 | 21 | X<sup>2</sup>=2.14 |
| RAVLT Immediate Recall<sup>f</sup> | 48.90 ± 11.0 | 48.35 ± 10.15 | t=0.81 |
| RAVLT Delayed Recall | 12.57 ± 2.2 | 13.01 ± 1.8 | t=0.25 |
| RAVLT Recognition | 6.49 ± 3.56 | 6.78 ± 3.32 | t=0.13 |

<sup>a</sup>Number of participants who had depression, taken from the Geriatric Depression Scale
<sup>b</sup>Number of participants who currently have post-traumatic stress disorder (PTSD)
<sup>c</sup>Number of participants who have had post-traumatic stress disorder (PTSD) in their lifetime
<sup>d</sup>Mini-Mental State Examination (MMSE) to check for cognitive impairment
<sup>e</sup>Number of participants who have at least one apolipoprotein E4 (APOE-e4) allele, indicating a risk of Alzheimer's
<sup>f</sup>The Rey Auditory Verbal Learning Test (RAVLT) is a neuropsychological assessment that measures a person's verbal memory
\*significantly different at p<0.05 (no significant differences observed here)

#### Adults with acute TBI:TRACK-TBI

Data from individuals with an acute mild TBI were obtained from TRACK-TBI ^15^. Full details of TRACK-TBI and participant inclusion criteria are found in supplementary materials 1. 461 participants aged 17-83 years with mild TBI (GCS 14-15; N= 374) and orthopedic controls (N= 87) were included. In-person outcome assessments were completed at two-weeks (including an MRI, RAVLT, and insomnia sleep index (ISI)), six-months (RAVLT and ISI), and 12-months post injury (RAVLT and ISI). Participant characteristics for TRACK-TBI are presented in Table 2.

**Table 2:** TRACK-TBI Participant Characteristics.

|  | **TBI (mean ± SD)** | **No TBI (mean ± SD)** | **Difference** |
| --- | --- | --- | --- |
| Age | 39.60 ± 15.83 | 39.58 ± 14.58 | t=-0.01 |
| Sex | 133 M, 241 F | 28 M, 57 F | X<sup>2</sup>=0.11 |
| Years of Education | 13.08 ± 2.5 | 13.19 ± 2.3 | t=0.39 |
| Household Income | \$35,000 to \$49,999 | \$35,000 to \$49,999 | t=0.77 |
| American Indian (%) | 1% | 1% | X<sup>2</sup>=0 |
| Asian (%) | 2% | 5% | X<sup>2</sup>=1.00 |
| Black/African American (%) | 20% | 11% | X<sup>2</sup>=3.42 |
| White (%) | 74% | 82% | X<sup>2</sup>=1.99 |
| More than one/Unknown (%) | 3% | 1% | X<sup>2</sup>=0.22 |
| ISI<sup>a</sup> 2 weeks | 9.51 ± 7.3 | 9.58 ± 5.9 | t=0.095 |
| ISI 6 Months | 7.69 ± 6.9 | 5.99 ± 5.3 | t=-2.49* |
| ISI 12 Months | 6.87 ± 6.6 | 6.79 ± 6.3 | t=-0.10 |

\*Indicates significant effect at p < 0.05.
<sup>a</sup>Insomnia Sleep Index (ISI)

### Standard Protocol Approvals, Registrations, and Patient Consents

The TRACK-TBI and ADNI study were approved by the institutional review board of each enrolling institution, and all participants or their legally authorized representatives completed written informed consent.

### Sleep assessments

The Pittsburgh Sleep Quality Index ^16^ was used to assess sleep disturbances in the ADNI/DoD sample. PSQI is a validated self-report questionnaire yielding information about overall sleep quality, latency, duration, efficiency, disturbances, medication use, and the effects on daytime function across a previous 30-day interval. The total score ranges from 0-21, with higher scores indicating worse sleep quality. PSQI has been used to assess insomnia in TBI and has been reported on previously in ADNI/DoD ^17^.

The Insomnia Severity Index (ISI) was used to assess sleep disturbances in the TRACK-TBI sample. ISI is a seven-item self-report scale that assesses problems related to sleep onset, maintenance, and awakening; dissatisfaction with sleep patterns; interference of sleep problems with daily functioning; and noticeable impairments and levels of distress due to sleep dysfunction in the last two weeks ^18^. The ISI has been assessed using TRACK-TBI previously^19^. The total score ranges from 0-28, with higher scores indicating worse insomnia.

### Memory assessments

The Rey Auditory Verbal Learning Test (RAVLT) was assessed in both ADNI/DoD and TRACK-TBI samples. RAVLT is a list-learning task measuring auditory verbal memory ^20^ and consists of five learning trials where 15 words are read to the participant and the participant is asked to orally recall as many as possible. After a 30-minute delay, the participants are asked to recall the original list of 15 words. Then, a recognition trial is administered, with 15 studied words and 15 non-studied words, where the participant determines which words were read from the original list. Outcome measures for RAVLT are total words recalled across five learning trials, recall following interference, recall following the delay, and the number of words correctly recognized with higher numbers indicating better memory performance.

### MRI acquisition

MRI acquisition details for ADNI/DoD are previously described ^21^. Briefly, MRI scans were acquired using 18 3T scanners and included a three-dimensional, T1-weighted scan (voxel size=1.2×1.0×1.0 mm^3^) and a DWI sequence featuring 5 b=0s/mm^2^ volumes and 41 diffusion-weighted volumes (voxel size=1.36×1.36×2.7mm^3^, b=1000 s/mm^2^).

MRI scans from TRACK-TBI were acquired across 11 centers using 13 3T scanners; standardization of the DWI measures across all 13 of the scanners was achieved using both a traveling volunteer and a diffusion phantom, ensuring that variability across scanners was negligible ^22^. Participants were scanned using a DWI sequence featuring 8 b=0s/mm^2^ volumes and 64 diffusion-weighted volumes at b=1300s/mm^2^ (voxel size=2.7×2.7×2.7mm^3^). Sagittal three-dimensional T1-weighted images were also acquired. All scans were performed within 2 weeks of injury.

### DWI preprocessing

Raw images were downloaded and used for this analysis. Preprocessing was performed using FMRIB Software Library (FSL) version 6.0. Data were motion- and eddy-current corrected. Skull-stripping was performed using the Brain Extraction Tool^23^. TOPUP ^24^ was used for correcting susceptibility-induced image distortions. The anatomical T1 scan was co-registered to the DWI scan using Advanced Normalization Tools (ANTs)^25^. DTIFIT ^26^ was used to fit a single tensor model at each voxel. Once these images were acquired, tract-based diffusion maps were calculated using TBSSv1.2 (Tract-Based Spatial Statistics)^27^. Each participant’s fractional anisotropy (FA) map was nonlinearly warped to Montreal Neurological Institute (MNI152) space (1mm^3^ voxel resolution) using the FMRIb58_FA template using FSL’s FNIRT^24^. The data of each participant were projected onto the skeleton to obtain axial diffusivity, defined as the diffusion coefficient along the direction of maximum diffusivity (i.e., the primary diffusion eigenvalue, λ1), reflecting changes in barriers along that direction ^28^.

Mean axial diffusivity was calculated across voxels within each of nine white-matter (WM) tracts-of-interest: left/right superior longitudinal fasciculus, fornix, left/right superior corona radiata, left/right anterior internal capsule, and left/right posterior internal capsule, as our pre-defined metric of axonal injury. The tracts were defined by mapping the Johns Hopkins University ICBM-DTI-81 White-Matter Labeled Atlas ^29^ regions to the WM skeleton in MNI152 space. A full list of these tracts and their projections can be found in Supplementary Material 2. We chose to focus on these specific tracts based on prior literature describing their roles in sleep-wake states and transitions and importance for memory formation and consolidation ^9,30^.

### Statistical Analyses

All statistical analyses were performed in RStudio Version 1.7. T-tests or Pearson’s chi squared test of proportions were conducted to test for differences in demographic variables between those with a TBI and those without.

In the ADNI/DoD sample, differences in average axial diffusivity upon study visit 1 (baseline) between groups (history of TBI vs no TBI) in each of the 9 *a priori* WM tracts-of-interest (Table 3) were assessed using general linear models with a main effect of group, controlling for MRI scanner site, age, education, race, depression, lifetime PTSD, current PTSD, APOE4 status, and total number of TBIs. Next, to test for the effect of axial diffusivity at baseline on sleep disturbances (PSQI), we ran separate (for each WM tract-of-interest) general linear models with a main effect of axial diffusivity and group (TBI, no TBI), controlling for the aforementioned covariates. We included an axial diffusivity by group interaction to test if the associations statistically differed by group. False discovery rate-corrected (FDR) marginal effects were then calculated for each model as a measure of the independent association of axial diffusivity at baseline on sleep separately in each group (TBI and no TBI). All *P* values were corrected for multiple comparisons using Benjamini and Hochberg’s false discovery rate (FDR) with a *q* level of 0.05, after pooling all *p* values from each model. To test for an effect of axial diffusivity at baseline on the change in memory scores (RAVLT Delayed Recall, Immediate Recall, and Recognition) over one-year (i.e., two timepoints: baseline and 12-months), we computed linear mixed effect models with fixed effects of axial diffusivity at baseline, time, an axial diffusivity at baseline by time interaction, and a participant-specific random intercept and slope; these analyses were performed in the TBI participants only, and were restricted to WM tracts that showed significant interaction effects in the generalized linear models.

**Table 3:** Axonal injury following acute mTBI and sleep over 1 year TRACK-TBI.

| Timepoint | *TBI Marginal Effect*<br>$\beta$, (95% CI), $p_{FDR-corrected}$ | *No TBI Marginal Effect*<br>$\beta$, (95% CI), $p_{FDR-corrected}$ | *Interaction $\beta$, (95% CI),*<br>*$p_{FDR-corrected}$* |
| --- | --- | --- | --- |
| Two weeks | 7.2e+04, (-1.4e+05, -5.0e+03), 0.04 | -2.1e+05, (5.8e+04, 3.6e+05), 0.01 | 2.827e+05, (-4.7e+05, -7.9e+04), $\leq .01$ |
| Six months | 1.6e+05, (-2.4e+05, -7.6e+04), $\leq .01$ | -2.1e+05, (-4.3e+04, 4.5e+05), 0.10 | 3.618e+05, (-6.7e+05, -1.4e+05), .04 |
| 12 months | 1.1e+05, (-1.9e+05, -8.5e+03), 0.03 | -1.1e+05, (-1.5e+05, 3.6e+06), 0.40 | 2.087e+05, (-5.4e+05, 1.3e+05), .23 |

To assess the generalizability of the ADNI/DoD finding, we tested the same hypotheses in an independent sample of participants with an acute (< two weeks) mild TBI from the TRACK-TBI dataset. To test for differences between groups (mTBI vs orthopedic control) in axial diffusivity, similar general linear models were performed with axial diffusivity in the ALIC (Figure 1) at two-weeks as the dependent variable, with a main effect of group while controlling for site, age, sex, education, and race (as previously reported^10^). To test for the effect of axial diffusivity two-weeks post-injury on sleep disturbances (ISI) over one-year (at two-weeks, six months, and 12 months), separate generalized linear models with a gamma family and inverse link function were run with a main effect of axial diffusivity at two-weeks and group (TBI vs orthopedic controls). An axial diffusivity by group interaction was again used to test if the associations statistically differed by group and FDR-corrected marginal effects were again used as a measure of the independent association of axial diffusivity at two-weeks on sleep separately in each group. Gamma generalized linear models were used owing to the heteroskedastic nature of the positively skewed outcome data. To test for the effect of axial diffusivity at two-weeks on change in memory (RAVLT Delayed Recall) over one-year (four time points), in the TBI participants only, a linear mixed effect model with fixed effects of axial diffusivity at two-weeks, time (two-weeks, six-months and twelve-months) and an axial diffusivity at two-weeks by time interaction, and a participant-specific random intercept and slope were computed, controlling for all covariates. Because TRACK-TBI also contained men and women with acute mTBI, we ran sex-specific interaction models to test for differences in these associations between men and women (Supplementary material 5). Significance was defined at p<0.05 for all models.

**Figure 1:**
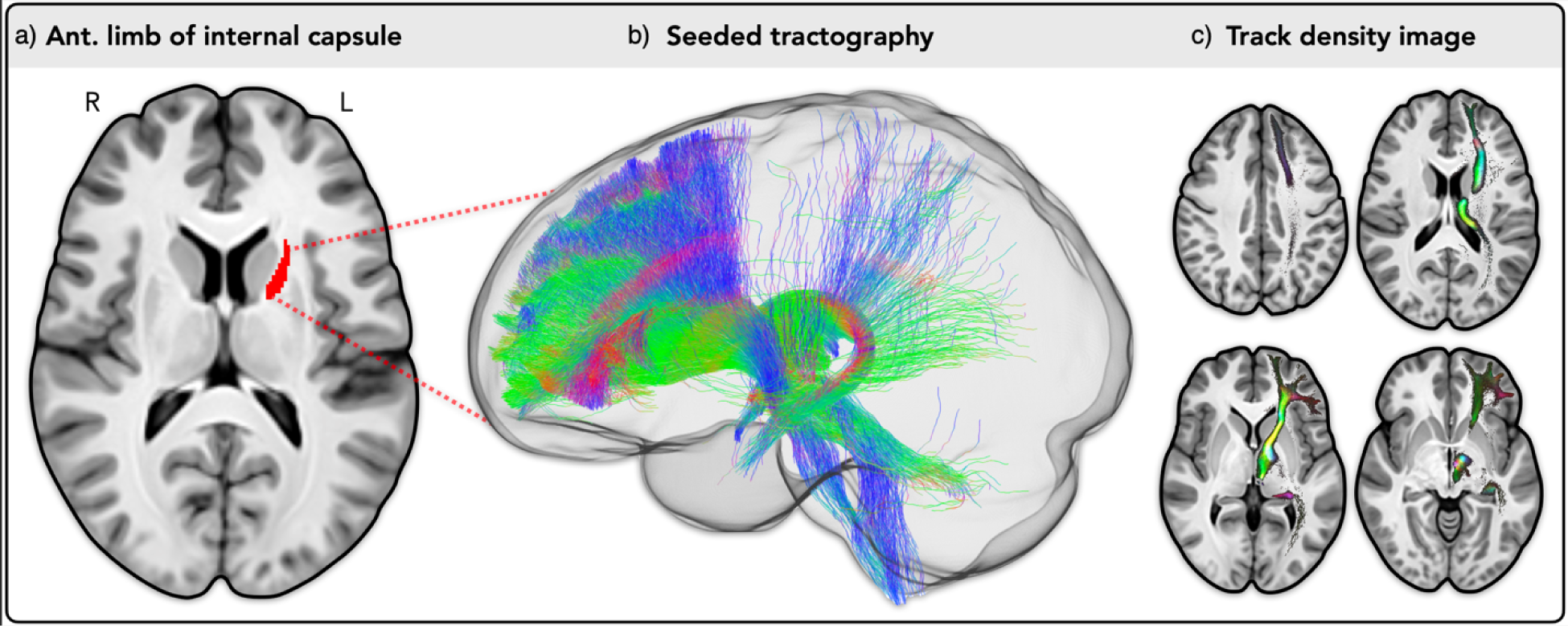
Typical projections of the left anterior internal capsule estimated from normative DWI data: a) left anterior internal capsule outlined by JHU ICBM-DTI-81. b) white-matter fibers seeded from the anterior limb of internal capsule using probabilistic tractography (20,000 streamlines), showing projections to the orbitofrontal cortex, ventromedial prefrontal cortex, dorsal anterior cingulate cortex, ventrolateral prefrontal cortex, thalamus, caudate nucleus, and the putamen. c) track density image of the streamlines seeded from the left anterior internal capsule, with brighter regions representing areas of higher fiber density and the color scale indicating fiber direction (green=anterior/posterior, blue=superior/inferior, and red=medial/lateral). These analyses were performed in MRtrix3 software with previously described methods ^31^, using an average fiber orientation distribution (FOD) template derived from healthy participant 3T DWI scans collected as part of the Human Connectome Project ^32^.

## Results

### Axonal Injury and sleep in older adults with a TBI history

Table 1 presents mean ± standard deviation values for all participant characteristics in the ADNI/DoD sample. No significant differences in demographic characteristics were found between those with a history of TBI and those without. No significant differences in axial diffusivity in any WM tract were found between those with a history of TBI and those without (Supplementary Material 3).

There was a significant positive relationship between baseline axial diffusivity in the left anterior internal capsule (Figure 1) and PSQI in the TBI group only [β = 9.52e+7, R^2^=0.06, 95% CI (1.5e+07, 1.4e+08), p_FDR-corrected_ ≤ .01] indicating that increased axial diffusivity was associated with increased sleep disturbances in those with a history of TBI. The association was statistically different between groups, where no significant association was found in those without a history of TBI [β = 0.09, R^2^=0.33, 95% CI (−1.2e+08, 9.6e+06), p_FDR-corrected_ = .21] (Figure 2). Full model results are presented in Supplementary Material 4.

**Figure 2:**
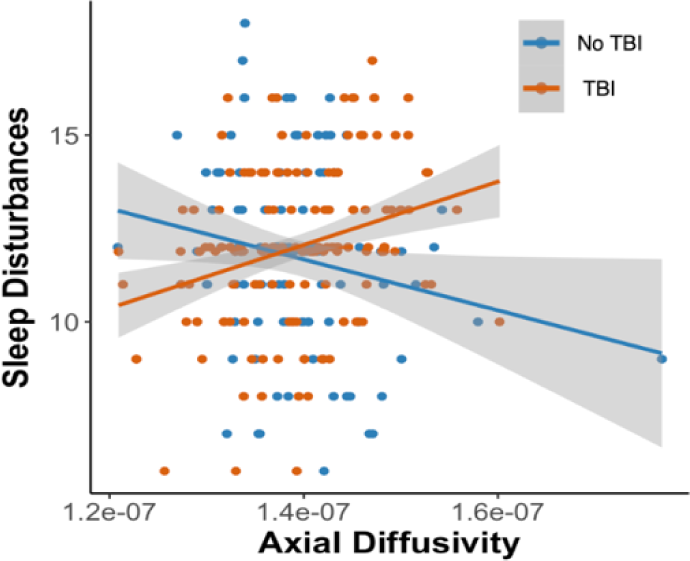
Scatter plot showing a significant positive association between axial diffusivity in the left anterior internal capsule (x axis) and sleep disturbances (as measured by the PSQI, on the y-axis) in the TBI group (orange slope). This association was significantly different to those without TBI, where the slope of the association was not significant (full model results are found in supplementary material 4).

### Axonal injury and cognition in older adults with a TBI history

A significant axial diffusivity by timepoint interaction was found for RAVLT delayed recall [β = −8.72e+7, 95% CI (−1.8e+08, −2.7e+07), η^2^= 0.04, p_FDR-corrected_ = .027], indicating that axial diffusivity in the left anterior internal capsule at session one was associated with changes in delayed recall over one year (Figure 3). Marginal effects revealed a significant effect of time when axial diffusivity was one standard deviation below the mean (Figure 3). No significant interaction was found for RAVLT recognition [p_FDR-corrected_ = .56] or RAVLT immediate recall [p_FDR-corrected_ = .97].

**Figure 3:**
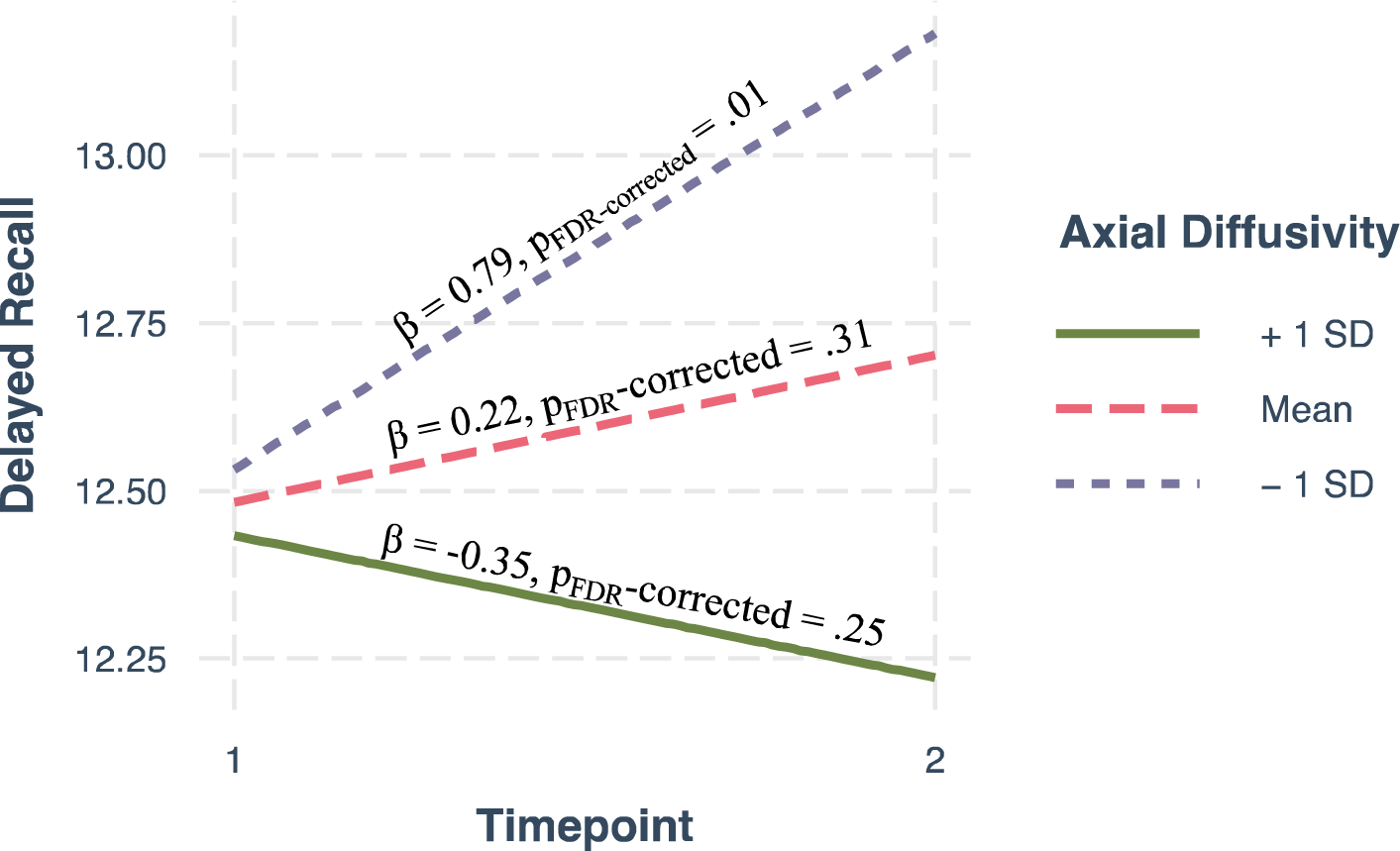
Marginal effects plot of axial diffusivity by time interaction showing that when axial diffusivity within the left anterior internal capsule is-1SD below the mean (less axonal injury) delayed recall memory (RAVLT) is better at one year. This association is opposite with axial diffusivity +1SD above the mean (more axonal injury) albeit not significant.

### Axonal injury following acute mTBI and sleep over one year

The TRACK-TBI sample showed significantly higher axial diffusivity in the left anterior internal capsule in the mTBI group compared to the orthopedic control group, after controlling for all covariates [β = 1.58e-08, 95% CI (5.3e-11, 3.2e-08), p = .037], as previously reported ^10,33^.

There was a significant positive relationship between two-week axial diffusivity in the left anterior internal capsule and ISI at two-weeks, six months, and 12 months post injury (Table 3) in the TBI group only, such that higher axial diffusivity acutely following mTBI was associated with higher ISI score (i.e., worse insomnia) at each time point. The association was statistically different compared to orthopedic controls at two-weeks and six-months, where no significant association was found in orthopedic controls at six-months or 12 months (Table 3 and Figure 4).

**Figure 4:**
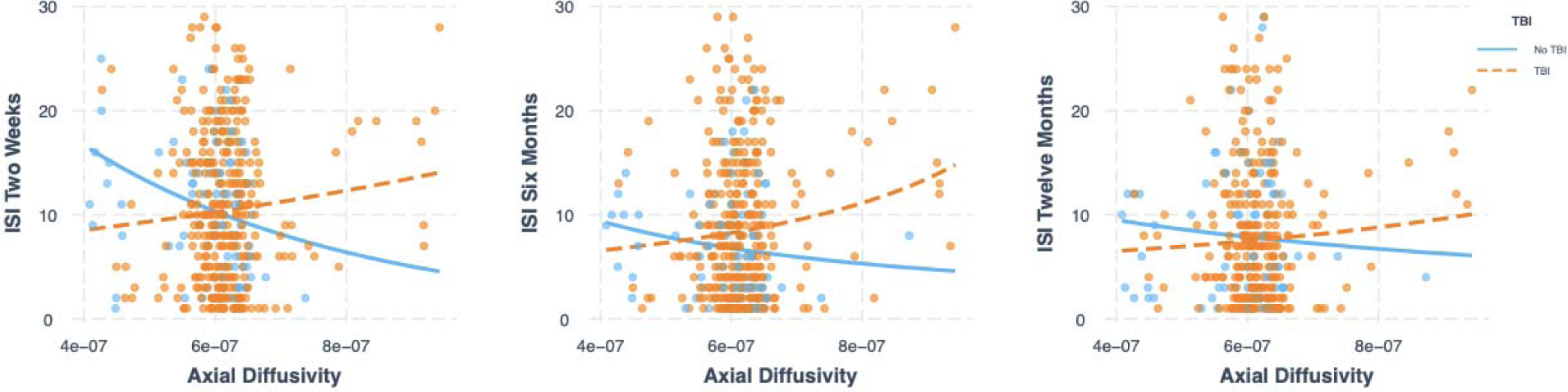
*S*catter plots showing a significant positive association between axial diffusivity in the left anterior internal capsule (x axis) at two-weeks post injury and sleep disturbances (as measured by the ISI, on the y-axis) at two weeks, six months and 12 months after injury in the TBI group (orange slope). This association was significantly different to those without TBI at two weeks and six months, where the slope of the association was not significant in the control group (at six months or 12 months).

### Acute axonal injury and cognition in mTBI

No significant *axial diffusivity* x *time* (two-weeks, six-months, twelve-months) interaction for RAVLT delayed recall was found [*p_FDR-corrected_* = 0.22], such that axonal injury to the left anterior internal capsule was not related to change in delayed recall over one year post injury.

## Discussion

In the ADNI/DoD sample of participants with a history of moderate to severe TBI, higher axial diffusivity in the left anterior internal capsule, thought to be a marker of axonal injury, was associated with more sleep disturbances. Axonal injury in this tract was further associated with longitudinal changes in delayed memory function. Using a second, independent sample of individuals with an acute mild TBI (TRACK-TBI data), we report robust evidence that axial diffusivity at two-weeks post-mTBI in the left anterior internal capsule was significantly associated with sleep disturbances at two-weeks, six-months, and twelve-months post-injury, demonstrating the generalizability of this finding across injury severities, populations and time since injury.

The anterior internal capsule carries both efferent and afferent thalamocortical projection fibers to many structures involved in sleep-wake states, including the thalamus, caudate nucleus, and putamen (Figure 4). Importantly, the thalamus has a strong influence on circadian rhythms and melatonin production ^34^, and is a key controller of arousal states during sleep-wake transitions ^35^. There is also evidence that thalamo-cortical network mechanisms are involved in memory formation ^35^. The caudate nucleus has been suggested to be a key structure involved in sleep-wake states due to its role in regulation of cortical excitability ^36^, with activity increasing during rapid eye movement (REM) sleep and decreasing during slow-wave sleep ^37^. If neurons are not firing properly in the caudate nucleus due to damage to the thalamocortical projection fibers, cortical excitability levels may be altered, potentially leading to disturbed REM and slow-wave sleep. The putamen is involved in autonomic regulation, with neuroimaging research showing decreased blood-oxygen-level-dependent (BOLD) signal activation after sleep deprivation ^38^ and smaller volume in sleep disorders ^39^. Hence, if the anterior internal capsule is damaged, projections to these important sleep-related structures may be dysregulated, leading to sleep-wake disturbances. Importantly, only the left, but not right, anterior internal capsule was associated with sleep disturbances. This however is in line with prior studies suggesting a left hemisphere dominance in the waking state, such that left hemisphere activity may be associated with fragmented sleep, greater sleep debt, and increased daytime sleepiness ^40^.

While definitive conclusions about directionality cannot be made, our findings from the TRACK-TBI data in individuals two-weeks out from their injury may suggest that 1) TBI leads to axonal injury in the left anterior internal capsule (as previously reported in Palacios et al., 2020) and 2) this damage to the left anterior internal capsule may have an acute impact on sleep that persists over time and up to twelve-months following the injury. Although a large body of literature suggests sleep quality may affect WM integrity over time ^9^, other evidence shows that damage to WM can precede sleep deficits. For example, multiple sclerosis, an auto-immune disease that causes demyelination, has been linked with the onset of sleep disorders ^11^, and the specific location of the WM lesions is associated with the type of sleep disorder ^11^. Similarly, in subcortical ischemic vascular dementia, which affects frontal-subcortical WM tracts, WM hyperintensity severity has been linked to sleep disturbances ^42^. Our results may be further supported by mouse models showing that myelin damage can lead to sleep impairments ^43^, suggesting that in certain cases, WM damage may be the cause of sleep impairments. Moreover, given the short delay between the injury and the DWI scan in the TRACK-TBI sample, it seems unlikely that poor sleep in those initial two weeks is solely responsible for the axonal injury^44^.

Nevertheless, these postulations about directionality cannot be extended to the older adult data. It is important to consider other factors that may have influenced the association between axonal injury and sleep disturbances in the ADNI/DoD sample, such as time since injury, comorbid PTSD, anxiety, depression, and numerous other factors that are associated with veterans. Nevertheless, our results showing that the associations are specific to those with a TBI demonstrate a robust link between axonal injury within the ALIC and sleep disturbances in TBI, irrespective of directionality.

The relative amount of damage to the left anterior internal capsule in those with a history of TBI influences longitudinal memory function. Specifically, less axonal injury was related to better performance on a delayed memory task across one year in the older adults with a history of TBI. Prior work has also shown associations between delayed recall and WM integrity in left anterior internal capsule in patients with schizophrenia^45^. The coupling of thalamocortical sleep oscillations with slow oscillations that emerge from the frontal cortices have been shown to be important for memory consolidation^46^. If lasting axonal injury to the anterior internal capsule from a TBI impacts the timely associations between these oscillatory mechanisms of memory consolidation, then this could provide one explanation as to why this measure of WM integrity was associated with changes in delayed memory over time. However, these results were limited to older adults in the ADNI/DoD sample with a history of TBI. The findings were not seen in the TRACK-TBI sample, for several possible reasons. The relationship between axonal injury and memory disturbances may not appear immediately after injury, and instead these disturbances may be a result of aging with this injury. Additionally, TRACK-TBI included a wider range of adults, with an average age of 39 years old, much younger than the ADNI/DoD sample and with mild TBI’s as opposed to moderate severe TBIs seen in the ADNI/DoD sample. Despite controlling for age in analyses, effects of aging on cognition may still be present. Additionally, the multiple timepoints for repeat testing over one year (more in the TRACK-TBI study) may have influenced performance due to practice effects. In the ADNI/DoD sample, we also found improved performance across testing instances that was also likely due to practice effects. Regardless, susceptibility to practice effects themselves have been posited to be an important early indicator of longitudinal cognitive decline^47^.

This study had limitations. First, while the sample across both datasets is large, many participants from the wider sample were not included due to not undergoing MRI or were excluded for incomplete data required for our analysis. Second, a history of TBI in the ADNI/DoD was self-reported, potentially leading to incorrect classification or missed TBIs from the control group. Third, the ADNI/DoD sample only consisted of male veterans and TRACK-TBI contained both men and women. Our supplementary analysis showed no effect of sex on the associations between axonal injury and sleep at baseline or twelve months but a differential effect at six months post injury. Given the importance of biological sex to the recovery of TBI, future research should study sex-specific research questions directly^48^. Fourth, TRACK-TBI used orthopedic controls, defined as individuals who had sustained an injury, but not to the head or neck; these other injury types may have transiently impacted sleep quality. This may explain why sleep disturbances at two-weeks post-injury were similar between the mTBI and orthopedic control groups, which resolved by month six in the control group only. Fifth, the sleep questionnaires were also self-reported, potentially introducing bias, although both PSQI and ISI are validated questionnaires to assess sleep. Additionally, there is no data on sleep complaints prior to injury, so causality cannot be directly inferred. The sleep quality questionnaires were different between datasets; however, this could be a strength of our study, suggesting our findings are reproducible across sleep measures. Seventh, ADNI/DoD included moderate-to-severe TBI, whilst TRACK-TBI included mTBI. Moderate-to-severe TBIs are expected to have longer consequences and associated focal injuries which may affect how generalizable these results are across groups. Eighth, the ADNI sample consisted of veterans with PTSD due to the nature of the ADNI study. PTSD may be a potential bias due to mixed effects of TBI and PTSD. However, PTSD was used as a covariate and there were no PTSD differences between groups. The TRACK-TBI sample did not have measures of PTSD, hence this bias could not be addressed here. We must take interpretations of these results with caution as this change in microstructure due to sleep disturbances may be accelerated by other comorbities, such as PTSD, anxiety, depression, among others.

## Conclusion

Using two independent datasets, this study provides unique insights into the acute and long-term associations of axonal injury and sleep disturbances in TBI. Our findings span multiple populations, ages, and times since injury, providing robust evidence of an association between axonal injury to the left anterior internal capsule and sleep disturbances and memory impairments in those with TBI. The results may suggest that quantification of axonal injury to the left anterior internal capsule could help clinicians identify patients at risk for continued sleep disturbances and related health consequences following TBI. Our results suggest that, if left untreated, this damage may portend poorer cognitive aging later in life. Clinically identifying axonal injury early after a TBI may provide a potential early therapeutic target to maintain WM integrity or improve sleep in TBI population. With the early implementation of such interventions, memory decline with age may be diminished in those with a prior TBI.

## Data Availability

Data was obtained from open-access databases LONI and FITBIR

## Acknowledgments

Data collection and sharing for this project was funded by the Alzheimer’s Disease Neuroimaging Initiative (ADNI) (National Institutes of Health Grant U01 AG024904) and DOD ADNI (Department of Defense award number W81XWH-12-2-0012). ADNI is funded by the National Institute on Aging, the National Institute of Biomedical Imaging and Bioengineering, and through generous contributions from the following: AbbVie, Alzheimer’s Association; Alzheimer’s Drug Discovery Foundation; Araclon Biotech; BioClinica, Inc.; Biogen; Bristol-Myers Squibb Company; CereSpir, Inc.; Cogstate; Eisai Inc.; Elan Pharmaceuticals, Inc.; Eli Lilly and Company; EuroImmun; F. Hoffmann-La Roche Ltd and its affiliated company Genentech, Inc.; Fujirebio; GE Healthcare; IXICO Ltd.; Janssen Alzheimer Immunotherapy Research & Development, LLC.; Johnson & Johnson Pharmaceutical Research & Development LLC.; Lumosity; Lundbeck; Merck & Co., Inc.; Meso Scale Diagnostics, LLC.; NeuroRx Research; Neurotrack Technologies; Novartis Pharmaceuticals Corporation; Pfizer Inc.; Piramal Imaging; Servier; Takeda Pharmaceutical Company; and Transition Therapeutics. The Canadian Institutes of Health Research is providing funds to support ADNI clinical sites in Canada. Private sector contributions are facilitated by the Foundation for the National Institutes of Health (www.fnih.org). The grantee organization is the Northern California Institute for Research and Education, and the study is coordinated by the Alzheimer’s Therapeutic Research Institute at the University of Southern California. ADNI data are disseminated by the Laboratory for Neuro Imaging at the University of Southern California. Data used in preparation of this article were obtained from the Alzheimer’s Disease Neuroimaging Initiative (ADNI) database (adni.loni.usc.edu). As such, the investigators within the ADNI contributed to the design and implementation of ADNI and/or provided data but did not participate in analysis or writing of this report. A complete listing of ADNI investigators can be found at: http://adni.loni.usc.edu/wp-content/uploads/how_to_apply/ADNI_Acknowledgement_List.pdf. Other data and/or research tools used in the preparation of this manuscript were obtained and analyzed from the controlled access datasets distributed from the DOD-and NIH-supported Federal Interagency Traumatic Brain Injury Research (FITBIR) Informatics Systems. FITBIR is a collaborative biomedical informatics system created by the Department of Defense and the National Institutes of Health to provide a national resource to support and accelerate research in TBI. Dataset identifier: NCT01565551. TRACT-TBI was funded by the US National Institutes of Health, National Institute of Neurologic Disorders and Stroke (grant U01 NS1365885) and supported by the US Department of Defense (grant W81XWH-14-2-0176). This manuscript reflects the views of the authors and may not reflect the opinions or views of the DOD, NIH, or of the Submitters submitting original data to FITBIR Informatics System.

## Author Contributions

ET: conception and design of the study, acquisition and analysis of data & drafting a significant portion of the manuscript or figures

GE: acquisition and analysis of data

AW: drafting a significant portion of the manuscript or figures LW: conception and design of the study

AS: conception and design of the study CH: conception and design of the study

TM: conception and design of the study, acquisition and analysis of data & drafting a significant portion of the manuscript or figures

