## Supplemental Materials for "Axonal Injury, Sleep Disturbances, and Memory following Traumatic Brain Injury"

**Supplementary materials**

1. Detailed participant inclusion and study design

ADNI/DoD

Data were obtained via a data use agreement through Laboratory of Neuro Imaging data image archive. ADNI began in 2003 by the National Institute of Aging and National Institute of Biomedical Imaging, led by principal investigator Michael W. Weiner. The primary goal of ADNI was to test whether MRI, positron emission tomography (PET), clinical and neuropsychological assessments could be combined to measure mild cognitive impairment progression and early Alzheimer’s Disease (AD) detection. Informed consent was obtained from all participants, and the protocol was approved by participating ADNI institutions. All participants provided written informed consent approved by local institutional review or ethics boards.

Participants were classified as having either a prior TBI or not using the RECTBIINJ.csv file (TBI Injury Questionnaire). These questionnaires determine when the injury occurred, its severity, and total number of injuries. The TBI was classified as moderate-to-severe based on the VA/DoD criteria for TBI with a loss of consciousness ≥ 24 hours, post-traumatic amentia > 24 hours, or alteration of consciousness ≥ 24 hours (Weiner et al., 2017). Moderate-severe TBIs occurred at a mean of 27 years of age. Depression and post-traumatic stress disorder (PTSD) were also characterized using the Geriatric Depression Scale (GDS) and the Clinician Administered PTSD Scale (CAPS). GDS scores of 0-4 are considered normal, 5-8 indicate mild depression, 9-11 indicate moderate depression, and 12-15 indicate severe depression, with interpretations dependent on age, education, and complaints. Current and lifetime PTSD was dichotomized with a score of 50 or above indicating PTSD.

TRACK-TBI

These data were downloaded from the open-access Federal Interagency Traumatic Brain Injury Research informatics system (FITBIR) via a data use agreement with Northeastern University. Participants were recruited from 11 academic level 1 trauma centers in the Unites States within 24 hours of injury, after evaluation in the emergency department or inpatient unit for TBI. Written informed consent was obtained, and the protocol was approved by the University of California, San Francisco and all enrollment site Institutional Review Boards. In-person outcome assessments were completed at two-weeks, six-months, and 12-months post injury. An additional assessment was carried out over the phone at three months. Data were collected in accordance with the TBI Common Data Elements (Maas et al., 2010). Assessments were administered and scored by trained study staff. Glasgow Coma Scale (GCS) classified injury severity upon arrival. The primary inclusion criteria for TRACK-TBI were patients who presented to the participating centers within 24 hours of injury with clinical indications for obtaining a computer tomography scan under the American College of Emergency Medicine/Center for Disease Control and Prevention Criteria (Jagoda et al., 2008). Inclusion and exclusion criteria for TRACK-TBI are reported in previous publications (Nelson et al., 2019; Yue et al., 2013). For our analysis, we included participants aged 17-83 years with mild TBI (GCS 13-15) and orthopedic controls that were recruited from the emergency room. Orthopedic injury causes included falls, pedestrian motor vehicle accidents, and bike accidents. These controls were excluded if a CT scan for suspicion of head trauma was required; if participants reported information such as loss of consciousness, amnesia, previous TBI, psychiatric, or neurological prevalent pathology; or if this study would be counterproductive for their sustained systemic injuries. Out of 568 participants from TRACK-TBI that had DWI scans, 461 had usable scans after preprocessing due to excess motion and brain, comprising 374 participants with TBI and 87 controls. Out of the 461 that had usable DWI scans, 10 participants did not report two-week sleep scores, 27 participants did not report three-month sleep scores, 11 participants did not report six-month sleep scores, and 66 participants did not report twelve-month sleep scores. Given the discrepancy in data collection method at the three-month assessment, we have excluded this time point from all further analyses.

2. Sleep tracts of interest

Table 1: *A priori* defined tracts-of-interest, their fiber types, and projections

| **ROI** | **Fiber Type** | **Projections** |
| --- | --- | --- |
| Anterior Internal Capsule | Projection Fibers | Comprises 4 regions: orbitofrontal cortex, ventromedial prefrontal cortex, dorsal anterior cingulate cortex, and ventrolateral prefrontal cortex. Includes bi-directional projections connecting the thalamus with the prefrontal cortex and the cingulate gyrus. It contains fiber tracts that travel between the caudate nucleus and the putamen. |
| Posterior Internal Capsule | Projection Fibers | The posterior limb is divided into two parts. The anterior half contains fibers of the corticospinal and corticobulbar tracts. The posterior part contains sensory neurons from the thalamus, fibers from the occipital lobe, acoustic fibers, and corticopontine fibers. |
| Superior Longitudinal fasciculus | Association fibers | Broken into three branches. The first branch connects the superior parietal lobule and precuneus with the superior frontal gyrus and anterior cingulate areas. The second branch originates in the posterolateral partial lobe and the angular gyrus and terminates in the dorsolateral prefrontal cortex. The third branch connects supramarginal gyrus to the inferior frontal gyrus. |
| Fornix | Association and commissural fibers | Broken into four parts. It connects the temporal lobe to the basal forebrain and diencephalon. It is a major output tract of the hippocampus. The posterior fibers continue through the hypothalamus and mammillary bodies to the anterior nuclei of thalamus. The anterior fibers end at the septal nuclei of the basal forebrain and nucleus accumbens. |
| Superior Corona Radiata | Projection Fibers | Cortical projections originate from pyramidal neurons from the motor cortex. It contains fibers from the corticobulbar, corticospinal, and corticopontine tracts. |

3. Between-group differences in tract-wise axial diffusivity in ADNI/DoD (no significant effects observed)

Table 2: Between-group differences in tract-wise axial diffusivity in ADNI/DoD

| **Axial Diffusivity (mm^2^/s)** | **TBI (mean ± SD)** | **No TBI (mean ± SD)** | ***p_FDR-corrected_* (95% CI)** |
| --- | --- | --- | --- |
| Anterior Internal Capsule (L) | 1.39e-07 ± 7.05e-09 | 1.39e-07 ± 7.05e-09s | .61 (-1.92e-09, 3.25e-09) |
| Anterior Internal Capsule (R) | 1.44e-07 ± 6.91e-09 | 1.43e-07 ± 7.77e-09s | .45 (-1.62e-09, 3.63e-09) |
| Posterior Internal Capsule (L) | 1.19e-07 ± 5.85e-09 | 1.17e-07 ± 5.54e-09s | .32 (-1.03e-09. 3.15e-09) |
| Posterior Internal Capsule (R) | 1.57e-07 ± 8.16e-09 | 1.58e-07 ± 7.08e-09s | .29 (4.2e+05, 1.6e+08) |
| Longitudinal fasciculus (L) | 1.66e-09 ± 1.03e-10 | 1.66e-09 ± 1.07e-10s | .63 (-4.76e-11, 2.86e-11) |
| Longitudinal fasciculus (R) | 2.39e-07 ± 1.09e-08 | 2.37e-07 ± 9.33e-09s | .89 (3.38e-09, 3.85e-09) |
| Fornix | 1.58e-08 ± 9.83e-10 | 1.58e-08 ± 8.69e-10 | .84 (-1.27e-09, 1.03e-09) |
| Superior corona radiata (L) | 1.00e-07 ± 7.91e-09 | 9.91e-08 ± 7.18e-09s | .07 (-2.24e-10, 5.32e-09) |
| Superior corona radiata (R) | 2.16e-07 ± 1.34e-08 | 2.14e-07 ± 1.25e-08s | .74 (-3.91e-09, 5.47e-09) |

4. Full model results of axial diffusivity in each ROI and PSQI in ADNI/DoD

Table 3: Full model results of axial diffusivity in each ROI and PSQI in ADNI/DoD

| **Axial Diffusivity (mm^2^/s)** | **Full model Adjusted R^2^** | **AD * PSQI interaction** (β, *p_FDR-corrected_* (95% CI)) | ***TBI marginal effect*** (β, *p_FDR-corrected_* (95% CI)) | ***No TBI marginal effect*** (β, *p_FDR-corrected_* (95% CI)) |
| --- | --- | --- | --- | --- |
| Anterior Internal Capsule (L) | 0.1890 | 1.207e-09, <.01 (6.8e+07, 2.2e+08)* | 95196285, (41121488.23, 149271082.80), 0.00 | -5096891, (-109292503.29, 7354520.09) , 0.09 |
| Anterior Internal Capsule (R) | 0.08414 | 6.406e-10, .22 (4.2e+05, 1.6e+08) | 60153324, (4553373.83, 115753275.03), 0.03 | -17837926.84, (-74821400.10, 39145546.41), 0.54 |
| Posterior Internal Capsule (L) | 0.03512 | -6.143e-11,  .51 (-4.6e+07, 1.6e+08) | 53662857, (-10456281.41, 117781995.41), 0.10 | -1819223.32, (-83686593.37, 80048146.73), 0.97 |
| Posterior Internal Capsule (R) | 0.09257 | 2.468e-10, .97 (-2.2e+09, 8.5e+09) | 17503122, (-28481549.13, 63487794.31), 0.45 | 18714763, (-44453861.16, 81883387.72), 0.56 |
| Longitudinal fasciculus (L) | 0.1026 | 3.167e-12, .51 (6.8e+07, 2.2e+08) | 2703035446, (-1383908.96, 74610629.08), 0.13 | - 440733323, (-36361681.00, 57034433.42), 0.84 |
| Longitudinal fasciculus (R) | 0.07164 | 1.836e-10, .59 (-3.3e+07, 8.6e+07) | 36613360, (-845412599.27, 6251483491.43), 0.06 | 10336376, (-4695015195.85, 3813548549.02), 0.66 |
| Fornix | 0.04893 | 1.013e-10, .51 (-5.9e+07, 3.1e+08) | 547996318, (169955579.27, 926037057.40), 0.00 | 41388316, (-454930501.56, 537707135.23), 0.87 |
| Superior corona radiata (L) | 0.07298 | 1.836e-10, .78 (-9.6e+07, 5.9e+07) | 104948, (-48698134.15, 48908030.18), 1.00 | 18588283, (-44281050.10, 81457616.99), 0.56 |
| Superior corona radiata (R) | 0.0766 | 7.551e-11, .78 (-3.6e+07, 5.5e+07) | 5438238, (-23661050.64, 34537527.62), 0.71 | -3967685, (-40340636.38, 32405265.95), 0.83 |

5. Sex differences in TRACK-TBI associations between axial diffusivity and insomnia sleep index

To test for the differential effect of axial diffusivity two-weeks post-injury on sleep disturbances (ISI) over one-year (two-weeks, six months, and 12 months) between men and women in TRACK-TBI, generalized linear models with a gamma family and inverse link function were run with a main effect of axial diffusivity at two-weeks and ISI at each timepoint by group interaction, controlling for all covariates. Post-estimation FDR-corrected simple slopes were calculated to test the independent association of axial diffusivity at two-weeks and ISI at each timepoint for each group.

Separate generalized linear models with the gamma family and inverse link function including an *axial diffusivity* (from the left anterior internal capsule at two weeks post-injury) x *gender* interaction term were performed with ISI total score at two-weeks, six months and 12 months after injury as dependent variables, controlling for all covariates (Supplementary Figure 2). No significant *axial diffusivity* x *gender* interaction was found for models two-weeks post injury [β=4.4e+04, SE= 8.9e+04, 95% CI (-1.40e+05, 2.1e+05), η^2^< 0.00, *p=* 0.62], six months post injury [β=2.5+04, SE= 1.1e+05, 95% CI -2.7e+05 1.9e+05), η^2^< 0.00, *p=* 0.82], or one year post injury [β =-3.6e+04, SE=1.5e+05, 95% CI -3.5e+05 2.6e+05), η^2^= 0.00, p = .82].

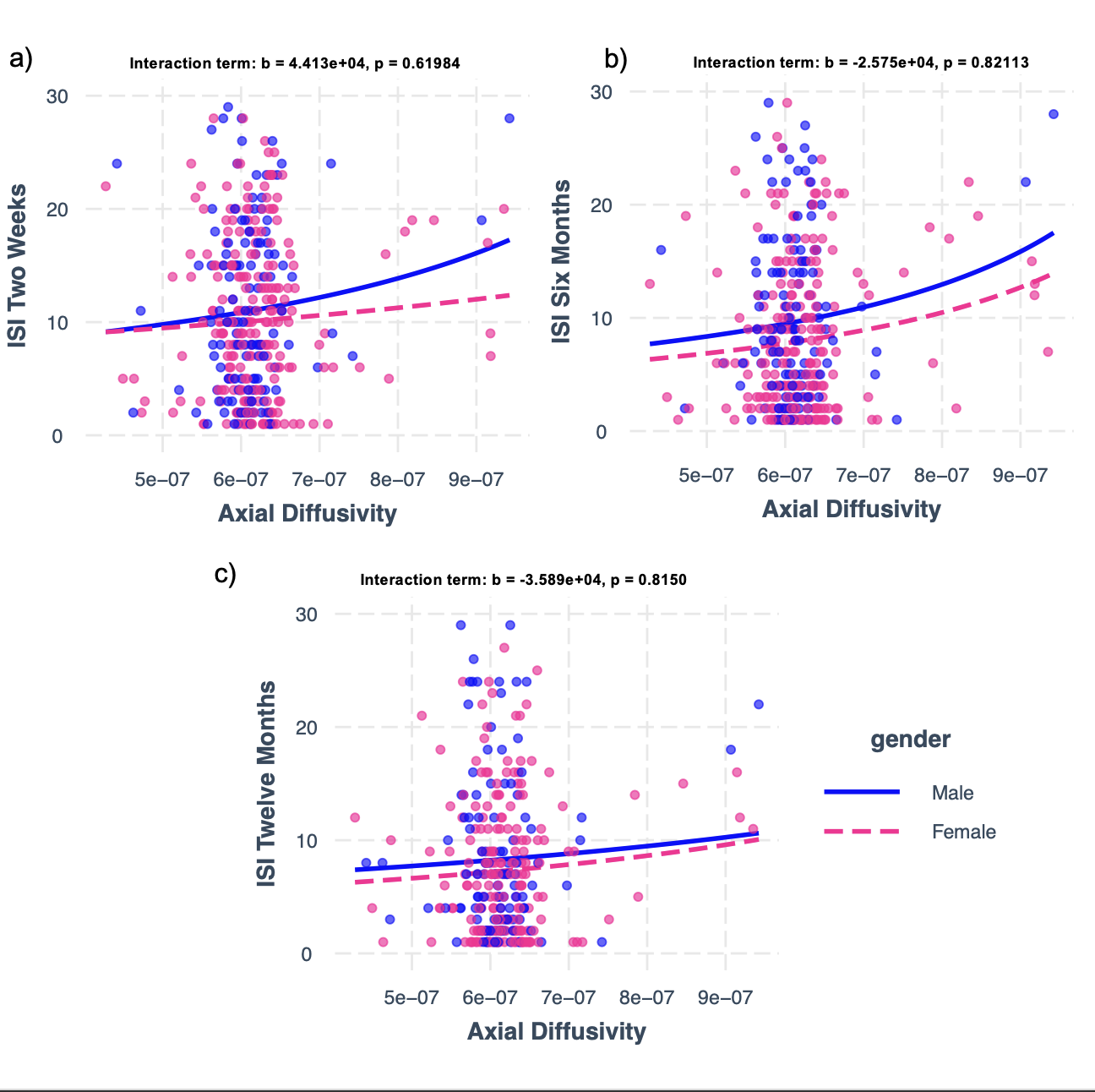

*Supplementary Figure 1:* General linear models with gamma family and inverse link function with a sex by ISI interaction on average axial diffusivity (measured acutely, <2 weeks post-injury) in the left anterior internal capsule at a) baseline, b) 6 months, c) 12 months
